# A Malaria Knowledge, Attitudes and Practice Survey in a Rural Community in Guinea

**DOI:** 10.1101/2021.06.18.21259155

**Authors:** Erin Holsted, Barclay Kadiebwe, Amer Sattar, Abigail Salthouse, Nirmal Ravi

## Abstract

Malaria is the top public health problem in the Republic of Guinea. In 2016, we conducted a cross-sectional household survey in Timbi-Touni, Guinea using community workers. The survey included respondent demographic characteristics, child health, child health promotion related to malaria knowledge, water and sanitation, and health services access. Majority of our respondents were women (89.41%) and had never been to school (71.18%). Slightly more than half the children were reported to have ever had malaria and 45% reported to have ever had diarrhea. We did not find any statistically significant association between gender or level of education and malaria knowledge. Eighty six percent of respondents had received a free bednet during national campaigns and 61% slept under a bednet the night before the survey. We found a statistically significant association between receiving information on malaria prevention and sleeping under a bednet. There was no statistically significant association between drinking water source and malaria or diarrhea. Both malaria and diarrhea were considered to be serious illnesses for adults and children by nearly all respondents. Insights from our detailed KAP survey can guide policy makers and practitioners who design and implement malaria control and prevention measures in Guinea.

## Introduction

Malaria is a life-threatening disease that poses a health threat to nearly half of the global population. The World Health Organization Africa Region continues to endure the largest burden of this illness. In 2018, 93% of malaria cases and 94% of malaria deaths occurred in sub-Saharan Africa (*World Malaria Report 2019*, n.d.). About 24 million children were estimated to be infected in 2018 in sub-Saharan Africa.

The Republic of Guinea is situated in the Gulf of Guinea in West Africa bordered by Sierra Leone, Liberia, Senegal, Guinea-Bissau, Mali, and Ivory Coast. Guinea’s population of approximately 13 million inhabitants are considered to be at risk of this vector borne illness that has year-round transmission with the peak transmission occurring during the July through October rainy season (*Guinea Malaria Operational Plan FY 2018*, n.d.). According to the National Strategic Plan of Guinea, malaria remains the number one public health problem accounting for a third of all patient visits (*Guinea Malaria Operational Plan FY 2018*, n.d.). The routine surveillance system in Guinea reported 992,146 cases of malaria and 867 deaths in 2016 (*Guinea Malaria Operational Plan FY 2018*, n.d.). Guinea is one of the countries with the highest percentage of severe anemia among children aged under 5 years who were positive for malaria, and it is the second highest leading cause of death for all age groups (*Guinea*, 2015; *World Malaria Report 2019*, n.d.). A recent national household survey of children aged 6 months to 9 years showed malaria prevalence between 44% and 61% with rural areas showing high prevalence (Beavogui, Delamou et al. 2020).

National Malaria Control Programs and donor agencies spent nearly $2.7B worldwide in 2018 for malaria control and eradication (*World Malaria Report 2019*, n.d.). In 2018, Guinea received $12M from The Global Fund and $15M from USAID (*World Malaria Report 2019*, n.d.). Guinea had two mass campaigns for distribution of Long-Lasting Insecticide-treated Nets (LLIN) in 2013 and 2016 (*Guinea Malaria Operational Plan FY 2018*, n.d.). LLINs are also given to pregnant women during prenatal care visits and 59% of the population had access to insecticide treated nets in 2016 (*World Malaria Report 2019*, n.d.). Guinea adopted a new National Strategic Plan in 2018 to reduce Malaria morbidity and mortality by 75% of 2016 level and achieve pre-elimination by 2022 (*Guinea Malaria Operational Plan FY 2018*, n.d.).

Despite such intensified efforts by donors and the National Malaria Control Program, the annual number of malaria cases continued to increase (*World Malaria Report 2019*, n.d.). National level data is extremely important for monitoring progress towards malaria elimination and informing public policy. However, local surveys can inform intervention design by providing the perspective of those facing the disease. Research of this kind is sparse in Guinea. However, previous studies in similar contexts have documented relationships between knowledge about malaria and several factors (Ahmed et al., 2009; Depina et al., 2019; Hlongwana et al.,2009). Low socioeconomic status and lack of education have been consistently associated with an overall lack of knowledge about malaria in the literature (Ahmed et al., 2009).

According to UNICEF, one-third of Guineans drink unsafe water which is the root cause of many health conditions and a large contributor to child mortality (*Water for Guinea*, n.d.). A 2020 study sought out to investigate if improved drinking water and sanitation conditions were associated with a decreased risk of malaria infection among children under the age of five in sub-Saharan Africa. Unprotected drinking water and lack of use of sanitation facilities were associated with increased malaria risks (Yang et al., 2020).

The purpose of this quantitative survey in a rural area of Guinea was to understand knowledge, attitudes, and practice (KAP) about malaria and to assess water and sanitation practices among community members. The information gathered can further inform and guide Guinea’s malaria control strategies towards the goal of pre-elimination by 2030.

## Methods

### The KAP Survey

A cross sectional household survey was conducted in 2016 in the sub-prefecture of Timbi-Touni, Pita Prefecture in the Republic of Guinea (See Figure 1). The questionnaire was administered to adult men and women who were present in the household when the interviewer visited. Investigators asked if the home visited had at least one child under the age of five. If the household did not have any child under five, the investigator thanked the individual and went to the next home. The interviews were done by community workers and those responsible for the local government health posts. They were trained to carry out the survey and spoke fluent French, as well as the local language Pular in order to communicate well with the participants. Community workers used mobile phones to record interview responses using an ODK (Open Data Kit) tool implemented by eHealth Africa.

**Figure 1.**
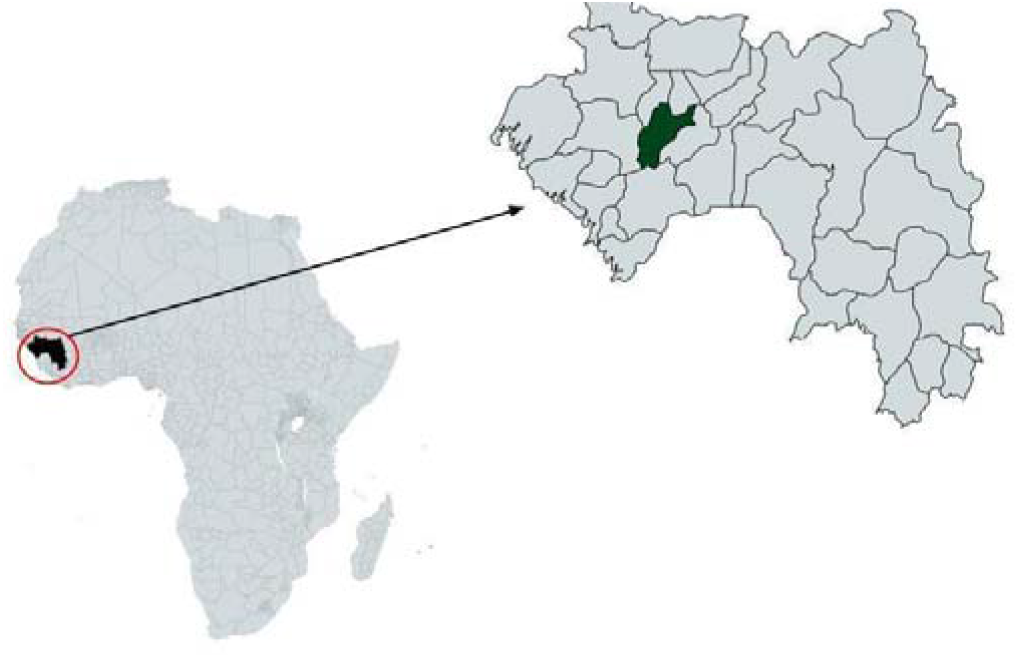
Guinea and the study site of Pita prefecture

In each village within each district, we visited the same number of households. For villages with a health facility, household No. 1 was chosen as the one closest to the health facility; for villages without a health facility, we considered proximity to the office of the village chief. Then, the following households were identified by a zigzag movement.

### Selection of variables

The questionnaire was structured to take approximately 50 minutes and was composed of 49 open-ended and multiple-choice questions. The interview was structured into six distinct sections: 1) respondent demographic characteristics, 2) child health, 3) child health promotion related to malaria knowledge, 4) water and sanitation, 5) health services access, 6) interviewee opinions concerning communication of malaria information and access to prevention methods.

### Data Analysis

Data analysis was done using SAS Version 9.4. Descriptive statistics were used to visualize the data using graphs created using Microsoft Excel. Chi-square test was used to determine differences between groups.

### Ethical Consideration

The interviewers attended a two-day training course in order to understand the questionnaires, master the collection tool, understand the informed consent script, understand the importance of ethics and confidentiality, and know the method of randomization.

The survey was only conducted after informed verbal consent of each participant was obtained. The questionnaire was anonymous and confidential. All responses were collected and stored electronically using password protected software.

Ethics approval was obtained from the Guinea National Ethics Committee for Health Research (96/CNERS/16).

## Results

### Study Population Characteristics

A total of 406 completed questionnaires from the interviewed households were used in the analysis of the survey data. A map of the health post locations that were surveyed is shown in Figure 2.

**Figure 2.**
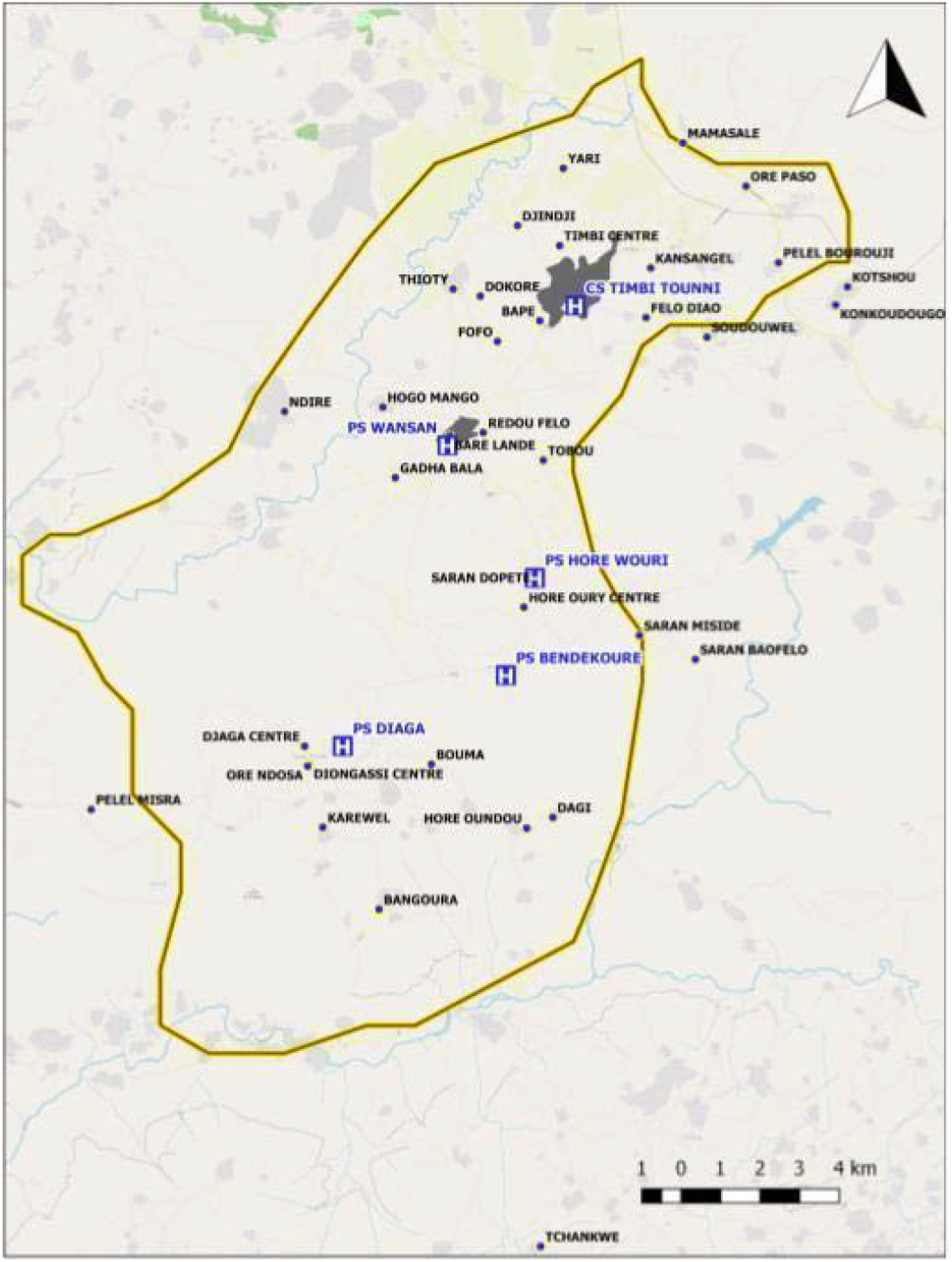
Timbi-Touni with surveyed villages

The majority of respondents were female (89.41%) and had never been to school (71.18%), followed by less than 6 years of education (22.17%), more than 10 years (5.42%), and more than 15 years (1.23%) of education. The average number of members in the household was 5.09±2.56. The average number of children under five in a household was 1.21±0.43.

The total number of children under the age of 5 included in this analysis is 493. The average age of the children was 28.83±17.36 months and 51.93% were female. Fifty four percent of the children in the survey were reported to have been born in a hospital.

### Child Health

The respondents, who were caregivers for the children in this survey, were asked if their children had ever had malaria or diarrhea. It was reported that 59.84% of children had ever had malaria and 45.44% of children had ever had diarrhea. Of the children who had ever had malaria, their caregivers reported that 52.54% of them slept under a bednet the previous night.

Additionally, the respondents were asked to report the last time that the child was taken to a health post and the reason for that clinical visit. A plurality of children were taken to a health facility in the past month (30.43%), followed by in the past six months (23.53%), past year (23.33%), past two weeks (20.69), and never (2.03%) as shown in figure 3. The primary reason for the clinical visit was to get a vaccine (36.31%). Other reasons were as follows: because the child was very ill (25.76%), for a routine checkup (25.56%), and other reasons (12.37%) (Figure 4).

**Figure 3.**
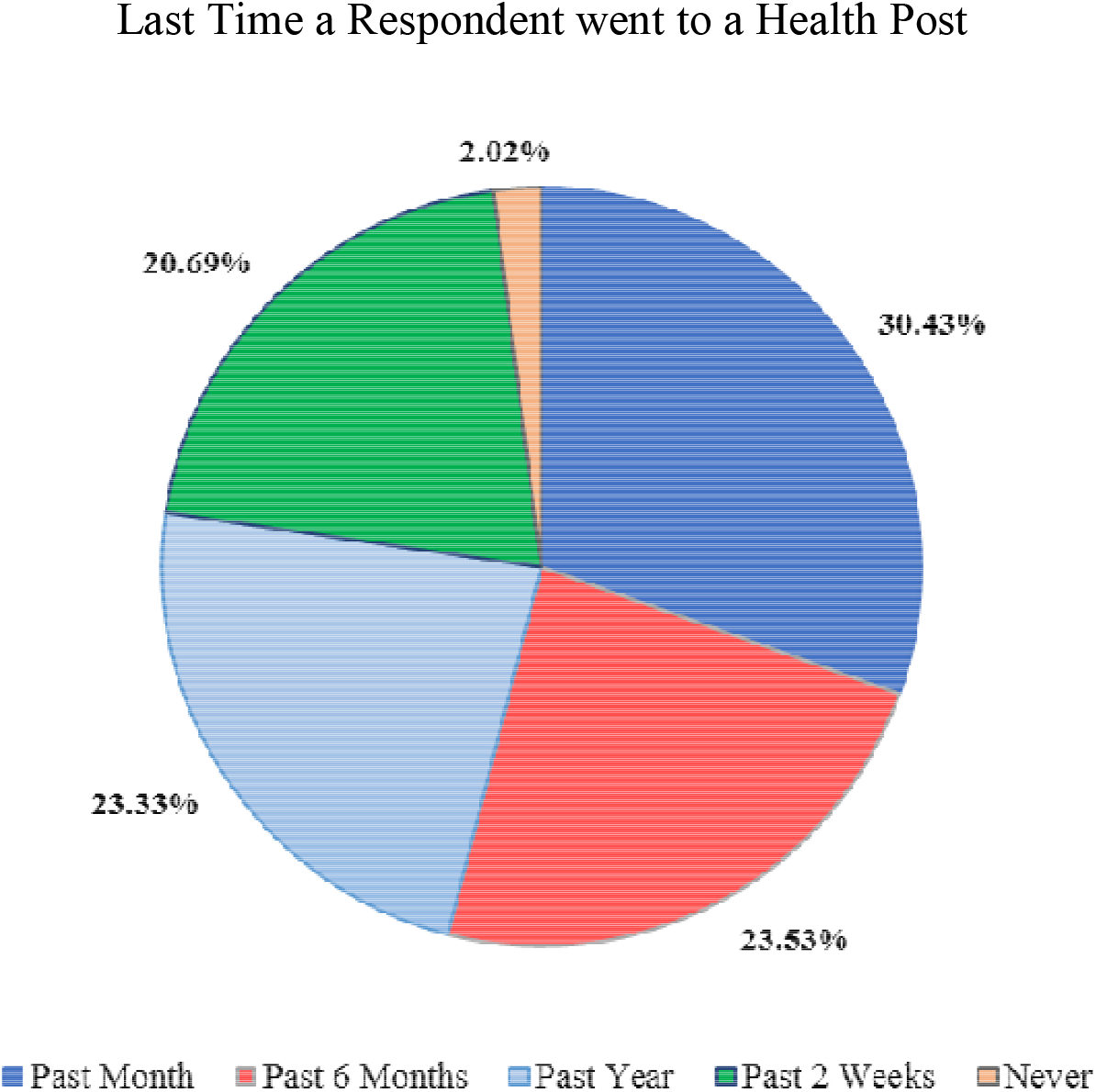
According to respondents, 30.43% went in the past month, 23.53% went in the last 6 months, 23.33% went in the last year, 20.69% went within the past 2 weeks, and 2.02% have never been to a health post.

**Figure 4.**
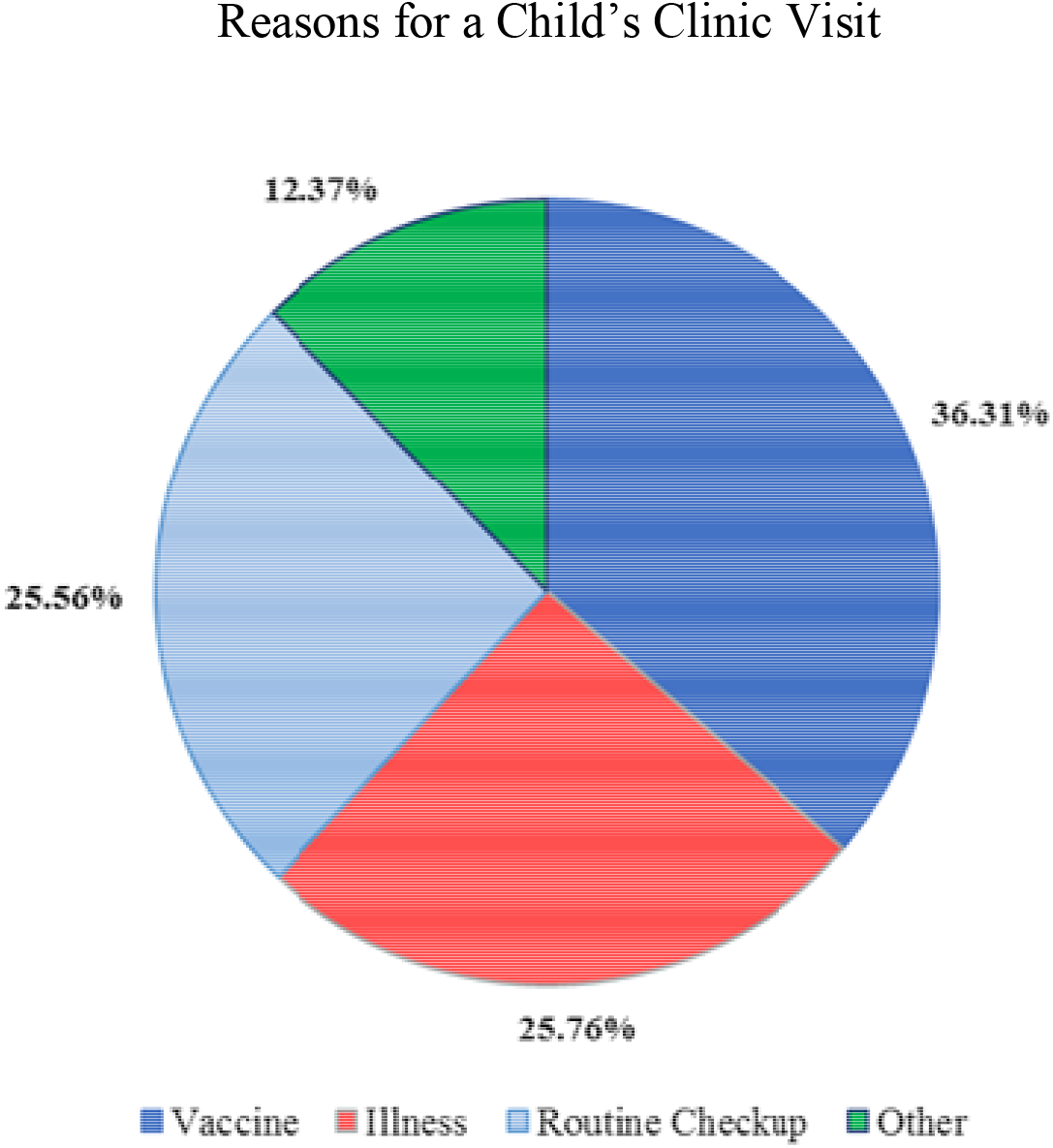
36.31% of respondents received a vaccine at their last health post visit, 25.76% went due to illness, 25.56% went to a routine checkup, and 12.37% went for other reasons.

### Knowledge of Malaria Transmission & Practices

Participants were asked to provide the common signs that an infant or child has malaria, and they were also asked to report in which way malaria was transmitted. These results are shown in Figures 5 and 6 respectively. The correct answer to the question “What are the causes of malariaã” served as the indicator for malaria knowledge. The majority of participants did mention that malaria was transmitted by mosquitoes (79.31%). However, only 39.66% correctly stated that mosquitoes were the only cause of malaria. There was no difference in the distribution of malaria knowledge between male and female respondents (p=0.1028). Additionally, there was also no difference in the distribution of knowledge of malaria amongst individuals who had completed various levels of education (p=0.3997).

**Figure 5.**
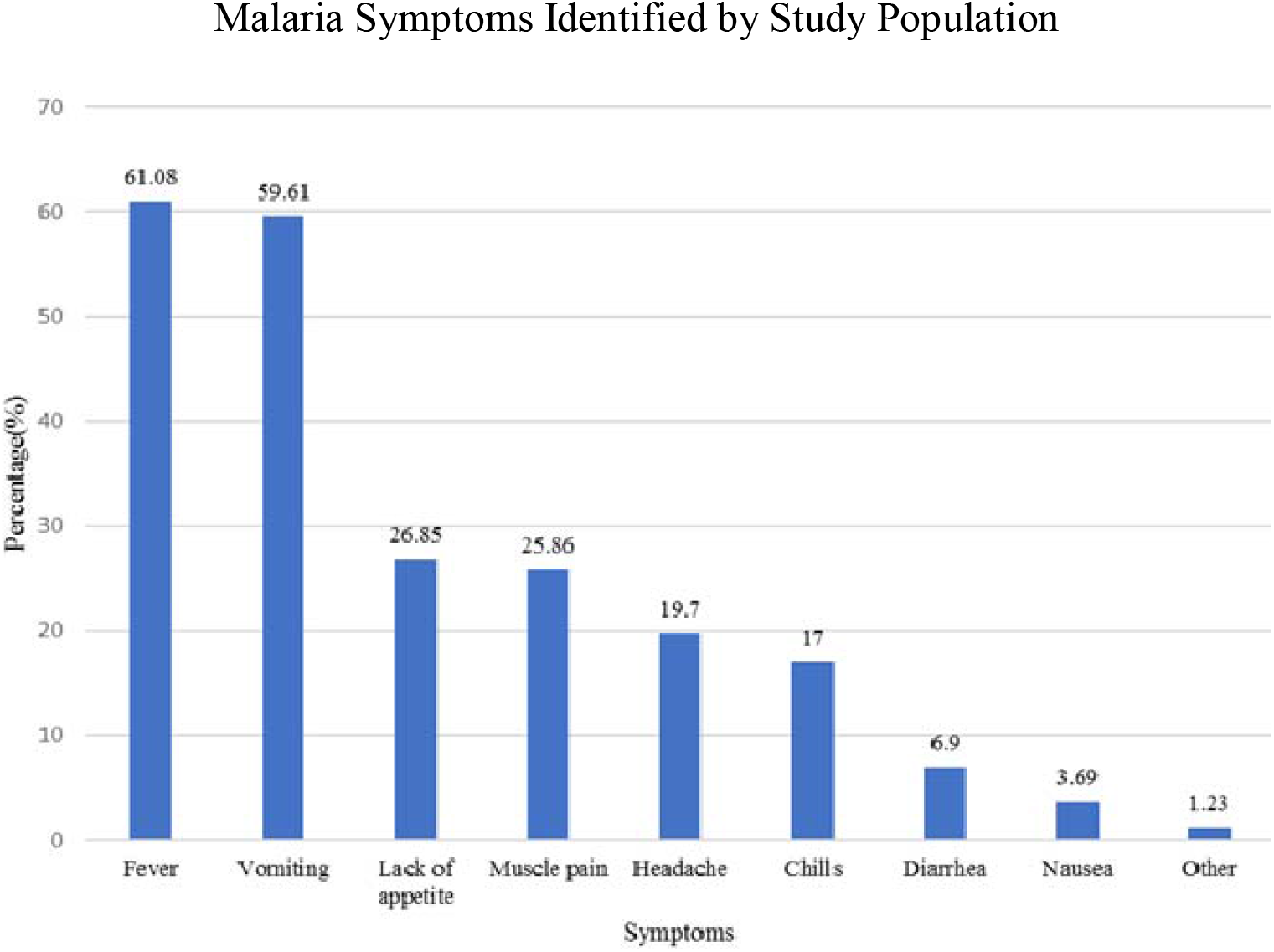
Common symptoms that an infant or child has malaria as identified included fever, vomiting, lack of appetite, muscle pain, headache, chills, diarrhea, nausea, and “other”.

**Figure 6.**
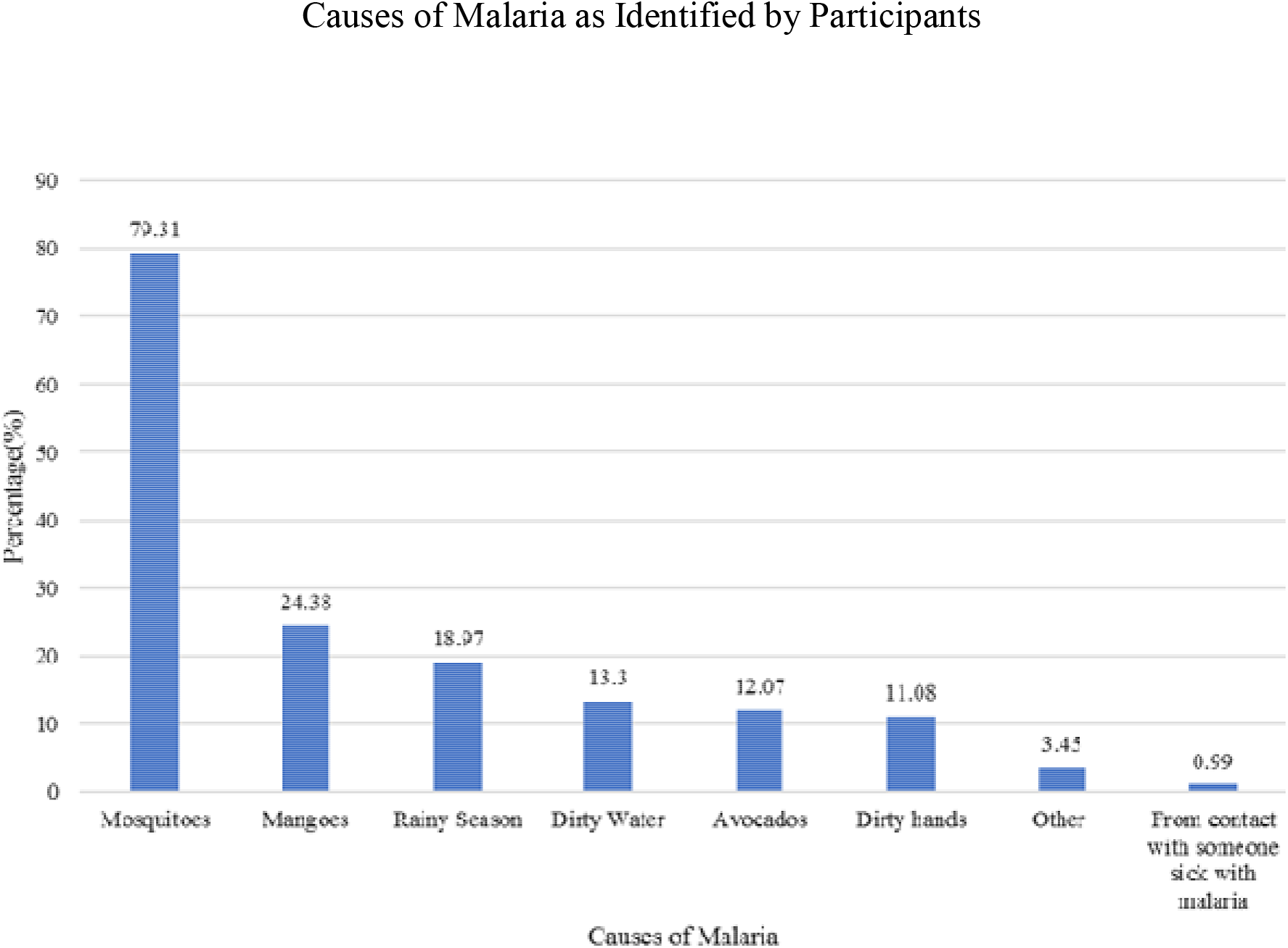
Causes of malaria as identified by the study population as mosquitos, mangoes, rainy season, dirty water, avocados, dirty hands, from contact with someone sick with malaria, or “other”.

With regards to malaria prevention practices, 85.71% of participants had received a bednet for free during a national campaign and 60.69% responded that they slept under a bednet the previous night. Sleeping under a bednet differed significantly between those who had and had not received information on malaria prevention (p=0.0014).

Sixty six percent of those who correctly identified the source of malaria slept under a bednet the previous night compared to 38.10% of those who did not correctly identify mosquitoes as the source of malaria (p=0.0001).

### Water, Sanitation, and Hygiene Practices

In order to collect information on people’s knowledge of childhood diarrhea, participants were asked to provide all the reasons they believed to be causes of diarrhea. A plurality of respondents attributed childhood diarrhea to dirty water (42.86%). Other reasons reported are shown in Figure 7. The majority of respondents indicated that their source of drinking water in the rainy season and the dry season was a borehole/hand pump, 63.55% and 60.59% respectively (Figure 8). Only 3.94% of individuals reported that they boiled water before consuming it. There was no statistically significant association between the source of drinking water during the rainy (p=0.49) or dry season (p=0.19) and malaria infection in children. Similarly, there was no statistically significant association between the source of drinking water during rainy (p=0.49) or dry season (p=0.84) and diarrhea in children.

**Figure 7.**
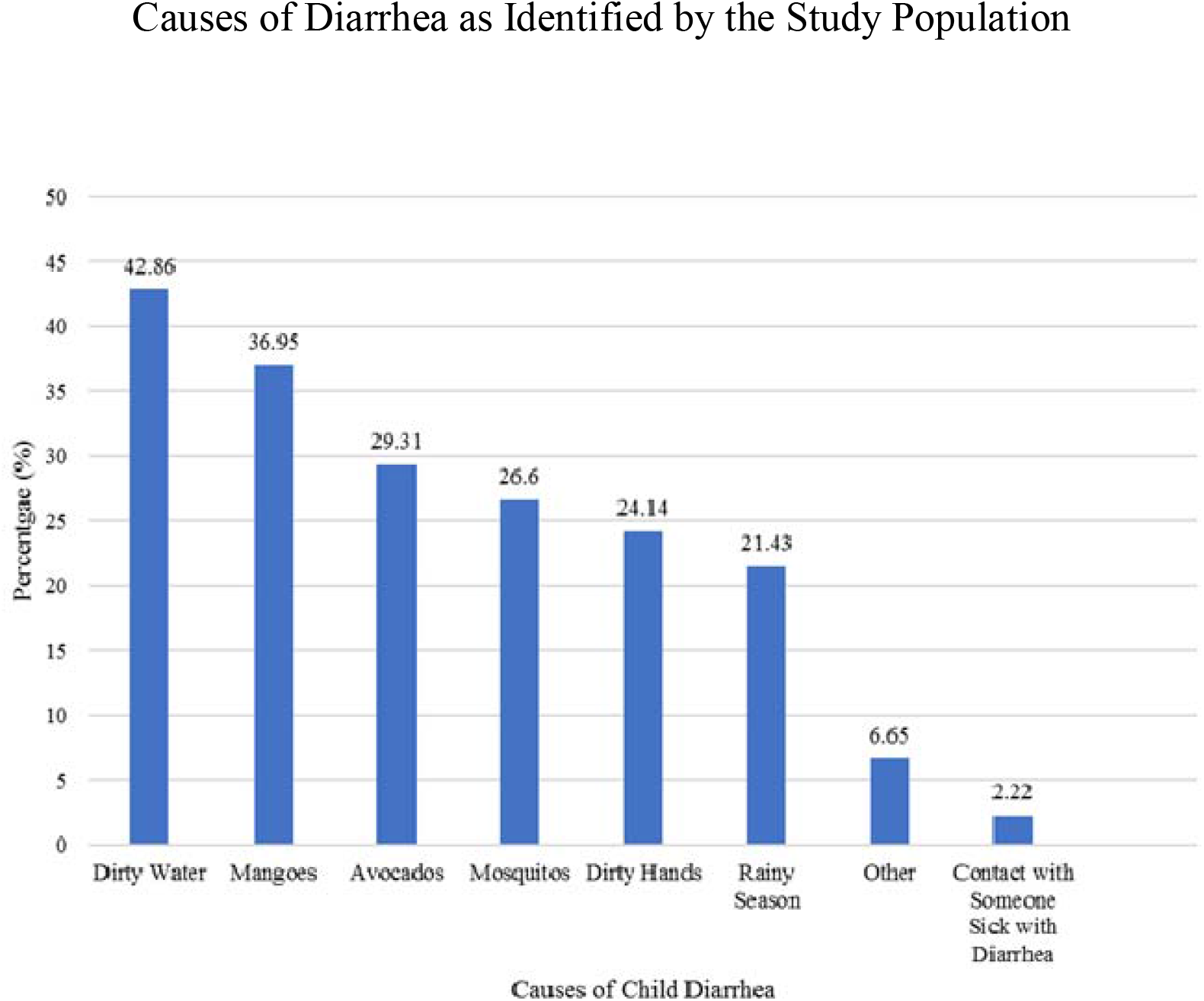
Causes of childhood diarrhea reported by the study population included dirty water, mangoes, avocados, mosquitos, dirty hands, rainy season, “other”, and contact with someone sick with diarrhea

**Figure 8.**
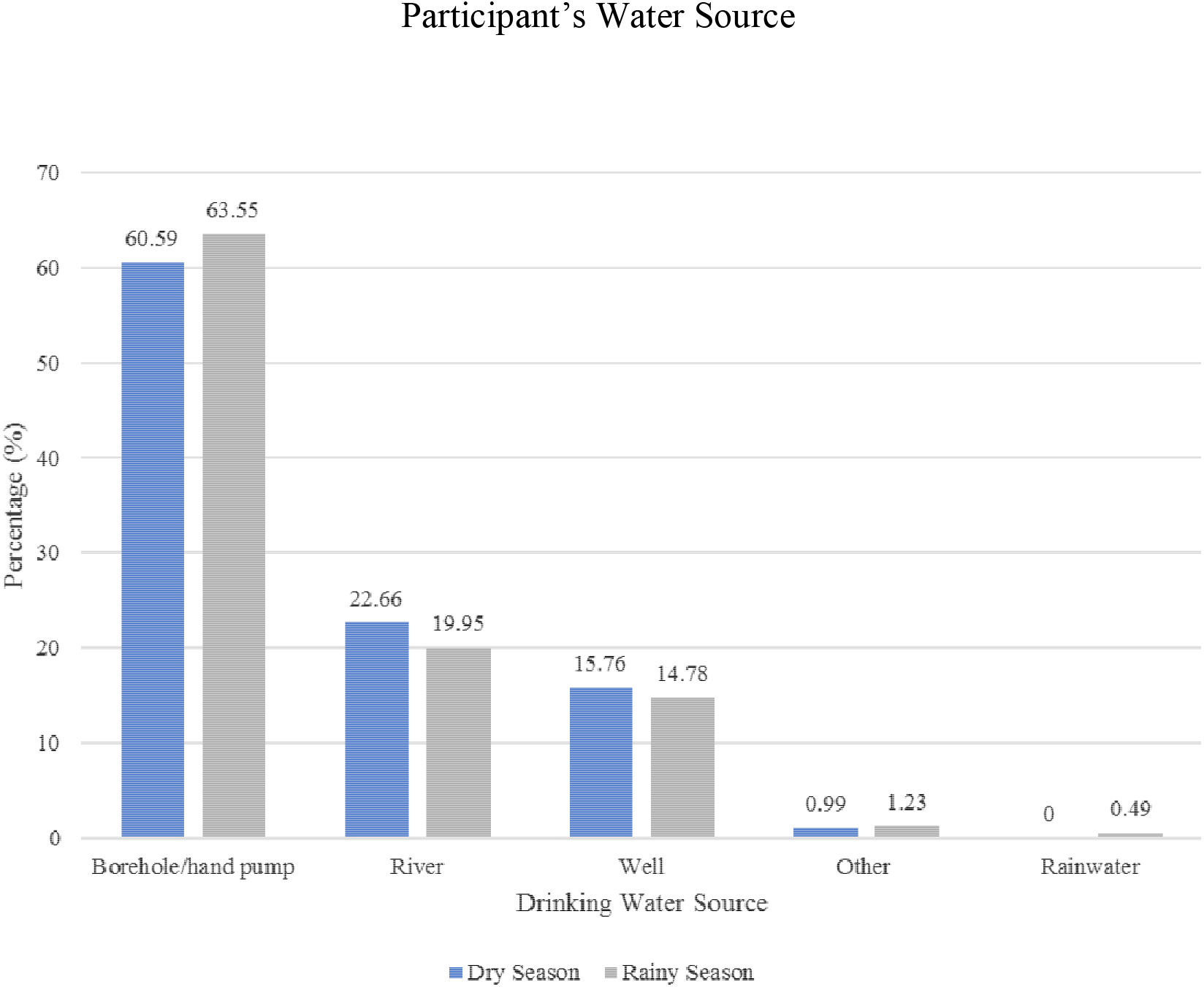
Drinking water sources were borehole/hand pump, river, well, other, or rainwater during dry or rainy season indicated by the study participant

The participants were also asked a series of questions regarding hand washing practices. On average, the respondents washed their hands 4.32 ± 4.40 times per day, mostly after using the toilet (43.35%), before eating (33.50%), and before praying (9.85%). The respondents were asked if they had soap in their household, and if so, they were asked to show the interviewer the soap. Ninety seven percent of the respondents said that they have soap in their household, however, only 74.94% of such individuals were able to produce the soap during the interview.

In our sample, for the children who ever had malaria, their caretakers washed their hands with soap on average four times per day. For the children who never had malaria, their caretakers washed their hands with soap on average five times per day. There was a statistically significant difference in caretaker hand washing frequency and their children having ever had malaria (p=0.0008).

Similarly, there was a statistically significant difference in caretaker hand washing frequency and their children having ever had diarrhea (p=0.046).

### Health Services Access

Interviewees were asked if they had any illness or injury in the past year that kept them from doing their usual work and other activities for at least one full day. If they answered yes to this question, they were asked a series of questions related to health seeking behavior. Twenty nine percent of respondents answered “yes” to having an illness or injury in the last year that prevented them from doing their usual activities. Of these individuals, 7.63% sought care from a traditional healer only, 42.37% sought care from the health post, and 39.83% accessed both forms of healthcare providers (Figure 9). 91.07% of those who accessed a traditional healer bought herbal medicines. The majority of individuals bought medicines from their health post (84.62%), compared to buying them at a pharmacy. Almost all respondents (94.95%) indicated that they were satisfied with the care they received at the health post or hospital they visited.

**Figure 9.**
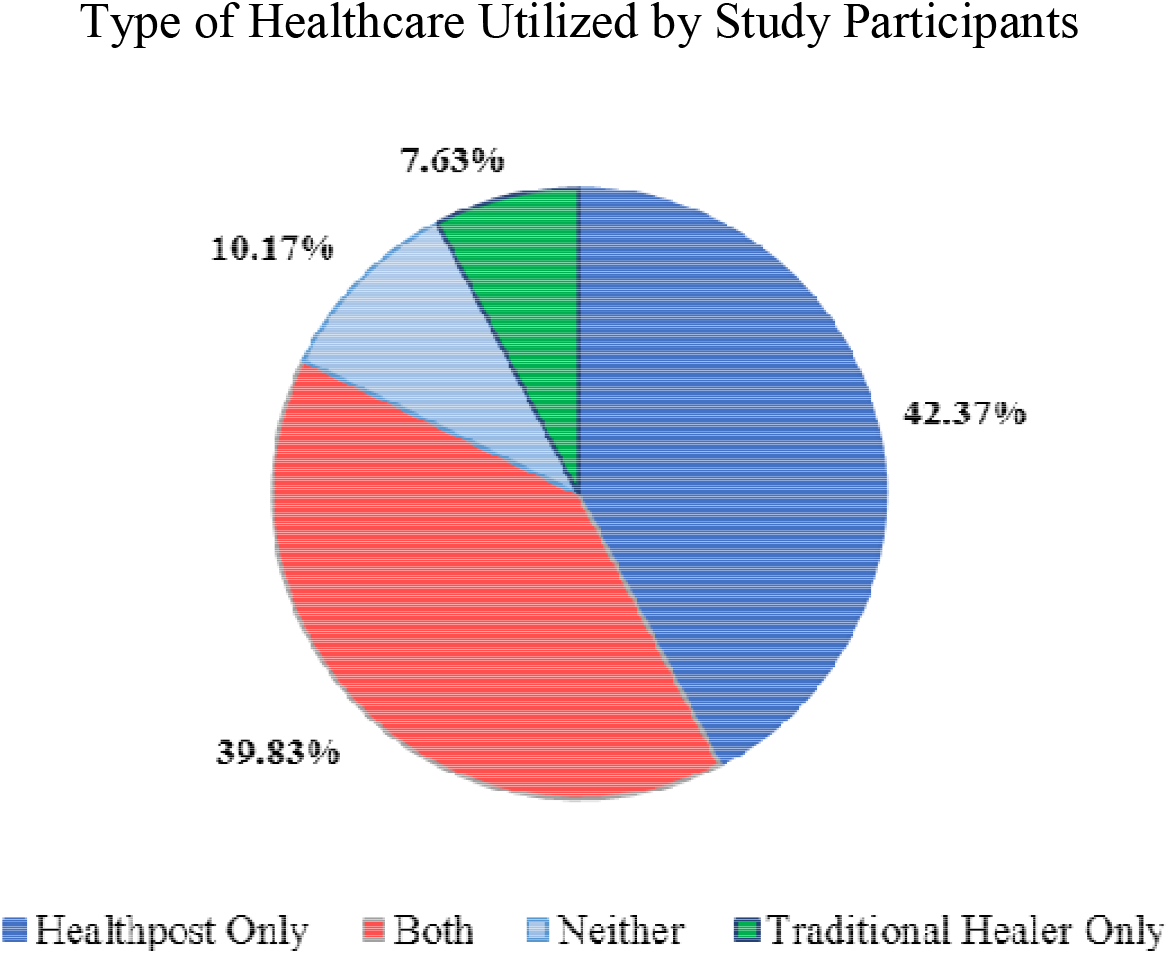
Type of health care provider included health post only, traditional healer only, both a health post and traditional healer, or neither.

All interviewees were asked about the cost of malaria treatment at their local health post. 30.05% said that it was free, 45.57% did not know the cost, and those who indicated it costed some amount of money reported an average of 76,786 Guinean Francs (GNF). Additionally, when asked to report the cost of a consultation with a clinician at a health post, 38.18% said that it was free, 48.28% did not know the cost, and those who indicated it cost some amount of money reported an average of 14,727 GNF. These results are shown in Figure 10.

**Figure 10.**
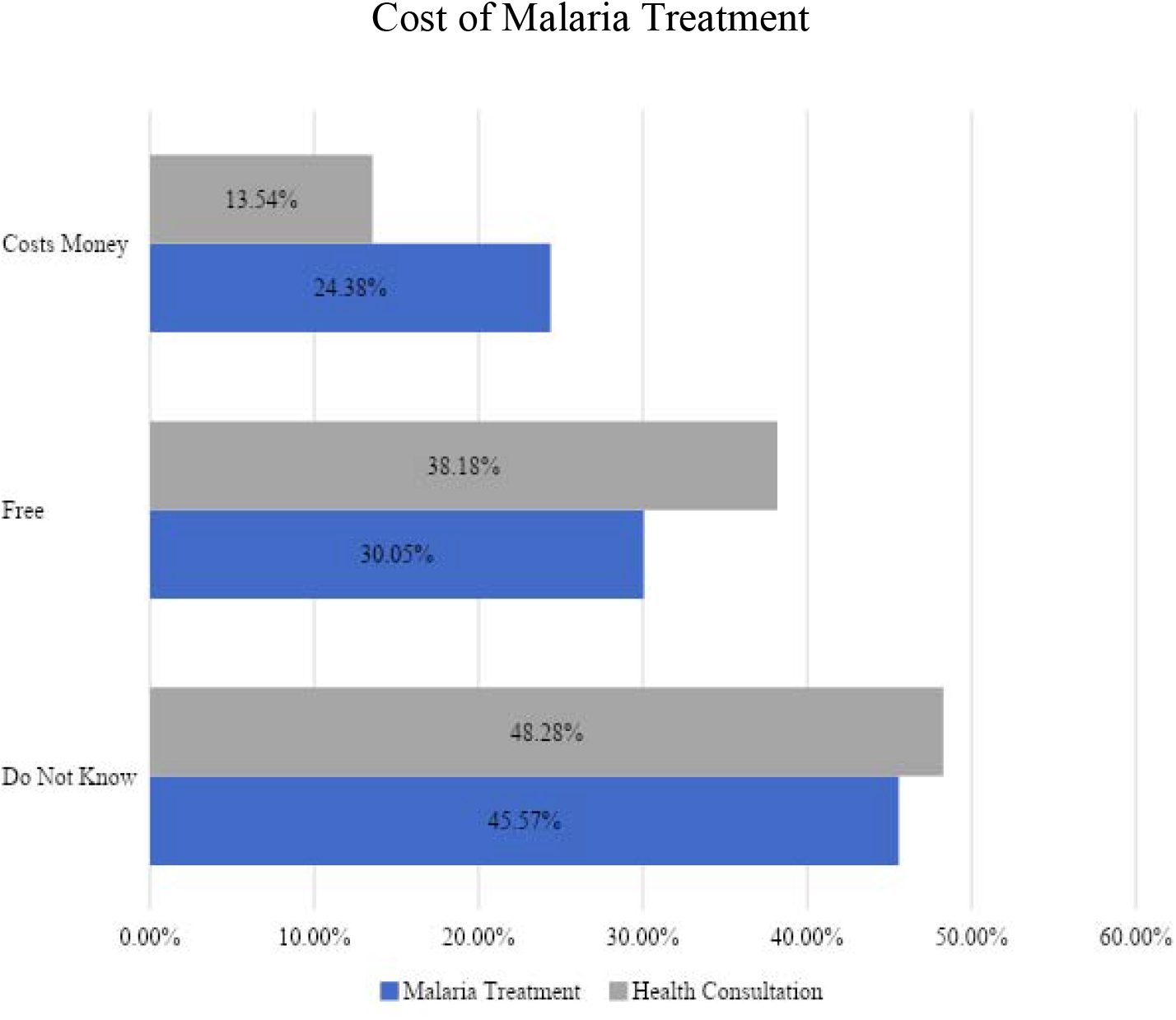
Cost of malaria treatment and health consultation

A large number of respondents, 91.63%, stated that there has been a time when they or someone else in their household was very ill and thought they should go to a health post but decided not to go for medical care. Over half the participants indicated that it takes at least 30 minutes to travel to a health facility, and 45.92% reported that it takes longer than 1 hour. With regards to those individuals who indicated that they had an illness or injury in the last year that inhibited them from doing usual activities, 39.32% reported that it took two days or more before they sought treatment or medicines after noticing the illness, followed by 46.15% who sought treatment the next day, and 14.53% the same day (Figure 11). This serves as an indicator for delay in seeking care.

**Figure 11.**
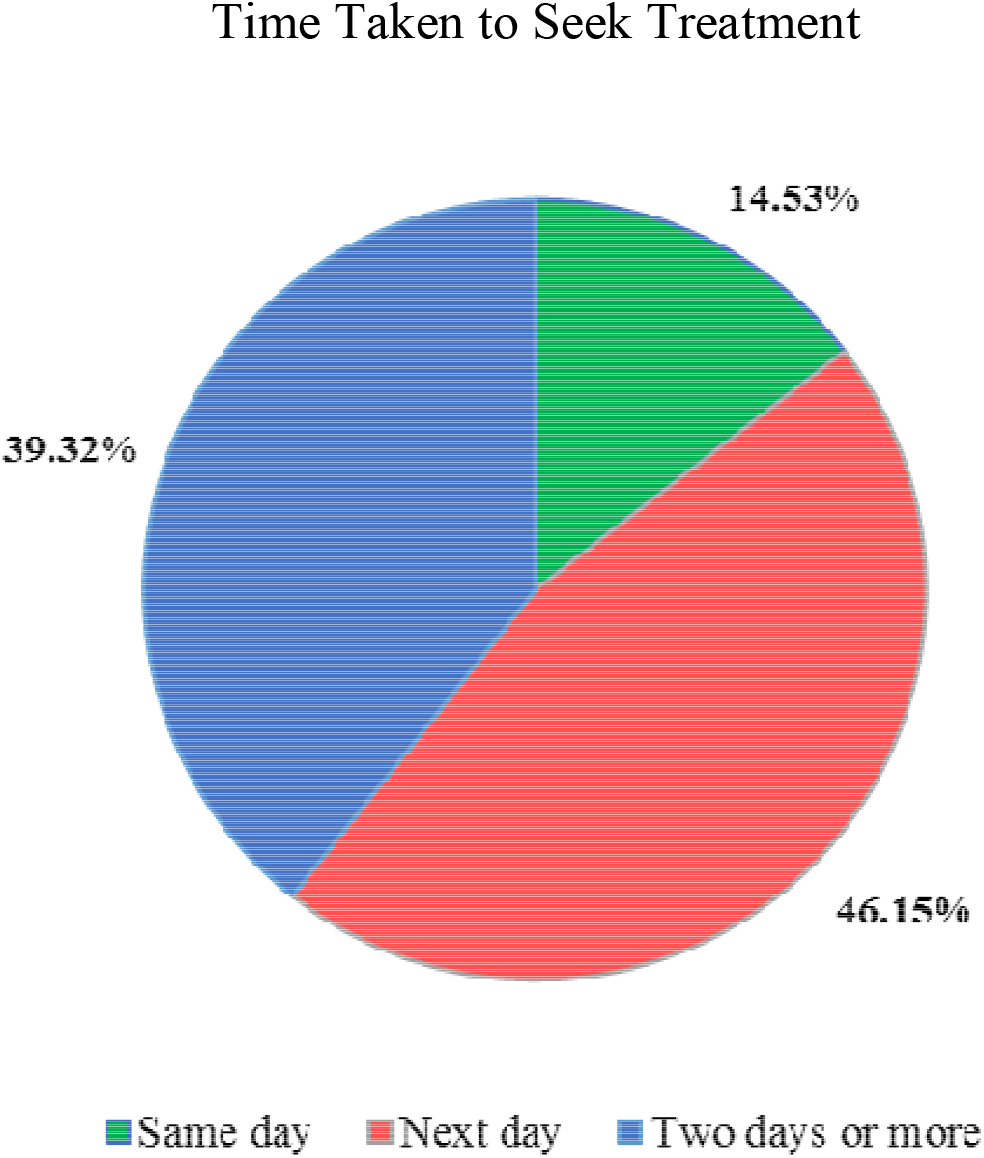
Time taken after the participant noticed his/her illness to seek treatment or medicines as same day, next day, or two or more days

### Disease Perception

A series of questions were asked to gain understanding of health-related opinions held by survey respondents.

### Illness

The vast majority of the respondents thought that both malaria and diarrhea were serious illnesses for adults and children. With regards to malaria, 98.28% of respondents thought it to be a serious illness in adults and 99.01% thought so for children. For diarrhea, 97.54% of respondents thought it to be a serious illness in adults, and 98.77% held this opinion in relation to children.

### Allopathic Medicine

As previously mentioned, many of the respondents in this survey sought the care of a traditional healer and bought herbal medicines. However, when they had malaria, 44.09% of individuals said they preferred pills as treatment, and 44.34% preferred an injection. For those that preferred an injection, it was for the following reasons: they are stronger (60.1%), they are faster (37.16%), and other reasons (2.75%).

### Health Information

Participants were asked to state their preferred and most trusted source of health information. Seventy-eight percent of individuals trusted radio for their health information. Other sources of information are shown in Figure 12.

**Figure 12.**
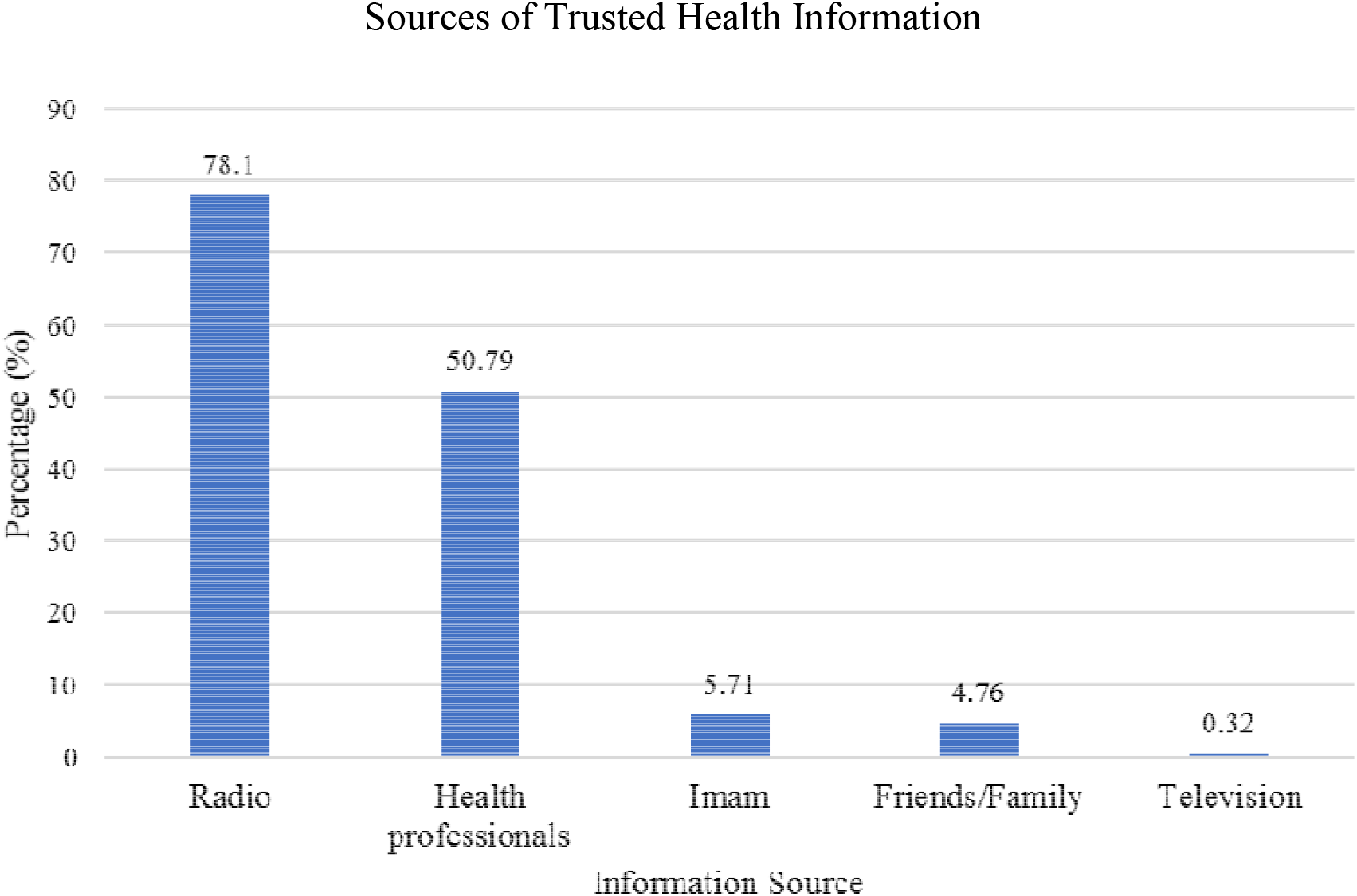
Trusted sources of health information include the radio, health professionals, Imam, friends/family, and television

## Discussion

Knowledge, attitude, and practice surveys are important for gaining insight into the misconceptions or misunderstandings that may serve as obstacles to prevention and treatment interventions that public health programs aim to implement (SPRING, 2014). It has been reported that childhood malaria prevalence is influenced by a complex interplay of socioeconomic, cultural, educational, geographical and environmental factors (Beiersmann, Sanou et al. 2007, Houeto and Deccache 2007, Clouston, Yukich et al. 2015). Because malaria is endemic to Guinea and all inhabitants are at risk of infection, it is imperative that local communities are assessed to identify gaps in knowledge so that resources and programs are allocated effectively. We noted an overall paucity of scientific literature on malaria in the Republic of Guinea, especially as it relates to water, sanitation, and hygiene practices.

### Caregiver level of education

Maternal education has been reported to be the best predictor of prompt malaria treatment in children under five in some countries in a multi-country analysis using national household survey data (Shah, Emina et al. 2015). In our survey, we found that three quarters of respondents had not attended any formal school. This percentage is very similar to what was reported from a different region of Guinea (Ruberto et al., 2014). This is much higher than what is reported from richer countries such as South Africa, Cabo Verde and Equatorial Guinea (Manana et al., 2015; DePina et al., 2019; Romay-Barja et al., 2015).

### Disease burden

We found almost half the children under the age of five in our survey to have had malaria and diarrhea in their lifetimes as reported by their caretakers. In contrast, a desk review of ministry of health records in Conakry revealed malaria prevalence less than 10% in children under five between 2009 and 2012 (Kouassi, de Souza et al. 2016). The difference could be due to the fact that our survey was in a rural area which tends to have higher malaria prevalence compared to urban areas. Malaria prevalence in children under 5 has been reported to be between 20% and 40% in countries as varied as Ghana, Democratic Republic of Congo, Uganda, and Burkina Faso (Tipke, Louis et al. 2009, Afoakwah, Deng et al. 2018, Mfueni, Devleesschauwer et al. 2018). These data show the enormity of the malaria disease burden among children under 5 in Sub-Saharan Africa.

We found that almost three quarters of the children had been taken to a health facility in the previous 6 months. More than half the child visits to a health facility were for preventive care such as routine checkups and vaccination. This is an encouraging finding that reveals the tremendous opportunity to provide routine preventive healthcare and health promotion counseling to young children and their caregivers by healthcare providers even in a rural setting.

### Caregiver gender

We did not find any significant difference in malaria knowledge between male and female caregivers, as well as different levels of education across all variables. Lack of significant difference in malaria knowledge between male and female caregivers could have been due to the low number of male caregivers in our survey. This contrasts with what has been reported from Bangladesh where level of education and gender were significant determinants of malaria knowledge and practice (Ahmed et al., 2009). This could be due to differences in education levels between the two study populations. Also, since radio was reported as the most trusted source of information in our population-which makes it more likely that everyone in the community had equal access to this information regardless of gender or level of education-it is conceivable that malaria knowledge was equally distributed among the study population.

### Knowledge of malaria

Almost four-fifths of those surveyed by us reported mosquitoes as one cause of malaria, although this proportion halved when using the more stringent criterion of only mosquitoes being the cause of malaria. This finding is similar to what has been reported from other countries (Hlongwana, Mabaso et al. 2009, Ndo et al., 2011). However, this high percentage in our survey was in contrast to the 19% reported by a 2014 survey done in a different region of Guinea (Ruberto et al., 2014). The high percentage of our survey respondents who had knowledge of the cause of malaria is a heartening sign since lack of knowledge was previously found to be a major barrier to malaria prevention (Maslove, Mnyusiwalla et al., 2009). Fever was the most common symptom associated with malaria in our study; this was similar to what has previously been reported (Romay-Barja et al., 2015).

### Bednet

A large percentage (86%) of the survey respondents had received a bednet during a national campaign. It is important to note that in our population, there was an association between receiving information on malaria prevention and sleeping under a bednet. A previous analysis of post-bednet-campaign surveys from Nigeria found significant increase in bednet usage after behavioral change communication through mass media (Kilian, Lawford et al. 2016). Secondary analysis of Nigerian Malaria indicator survey showed that topic-specific social and behavioral change communication improved the use of ITNs among children (Zalisk, Herrera et al. 2019). This information suggests that distributing long-lasting insecticide-treated mosquito nets through mass campaigns should be accompanied by an educational component. Distribution of free bednets has also been shown to increase its usage and decrease inequity in bednet ownership in Northern Nigeria (Ye, Patton et al. 2012). Bednet ownership was 55% and their use was 79% in a survey from Forecariah district of Guinea (Ruberto et al., 2014). The difference in ownership and use of bednets from the two Guinea surveys could be attributed to the study year, geography, and culture. Our survey was done five years after the survey by Ruberto *et al*, and there were two LLIN mass campaigns in the years intervening the two surveys. Interestingly in South Africa, a country that was in the malaria elimination phase in 2016, 99% of respondents correctly identified the cause of malaria, but bednet use was reported as 2% during a survey (Manana et al., 2017). Cabo Verde-another elimination country where 88% of survey respondents knew the cause of malaria and 97% had heard of mosquito nets-also showed a low 19% bednet use (Depina et al. 2019). It is possible that as malaria prevalence decreases and countries move towards elimination, fewer people use bednets as a method of malaria prevention. A study from Ghana, a richer country compared to Guinea, found no significant association between ITN use and malaria infection among children, except among children whose mothers had at least a secondary education (Afoakwah, Deng et al. 2018). Television was also found to be the best strategy to convey malaria education in Ghana (Afoakwah et al., 2018). These differences between countries show that the most effective malaria prevention and control approaches vary depending on local malaria prevalence (DePina et al., 2019). Because it is often reported, and supported by the data in our study, that individuals access health information through radio and television, malaria education can be effectively delivered through these platforms (Ahmed et al., 2009; DePina et al., 2019). We found a statistically significant association between correctly identifying mosquitoes as the source of malaria and use of bednet the night before the survey. Since bednet usage is a major intervention to decrease malaria incidence, our finding underscores the importance of malaria knowledge in improving bednet usage and thus reducing its burden in Guinea.

### Water, Sanitation and Hygiene

A recent study concluded that water and sanitation conditions are associated with the risk of malaria among children under five years old, and the evidence we have supports these findings (Yang et al., 2020). For participants in our survey, there was a statistically significant difference in caretaker hand washing frequency between children that ever had and had not had malaria. This relationship was also seen for reported diarrheal illness. It would be interesting to further investigate the relationship between drinking water source and malaria risk because the majority of our survey participants obtained their water from a borehole/hand pump. These are relatively clean sources of water that do not create stagnant water areas for mosquitos to thrive nearby.

### Healthcare access and practices

Approximately 40% of our respondents sought care from government health posts while 40% sought care from both government health posts as well as traditional healers. In rural Burkina Faso, 72% of children with malaria received modern treatment while 18% received traditional treatment (Tipke, Louis et al. 2009). Local perception of disease was also found to affect treatment choice in Burkina Faso (Beiersmann, Sanou et al. 2007). A similar number of respondents sought care from village doctors and drugstores in Bangladesh, which demonstrates the important roles played by traditional and informal healthcare providers in diverse developing countries (Ahmed et al., 2009).

Our survey found that more than half the respondents thought malaria treatment or consultations were not free at the level of the government health posts. This is a cause for concern as malaria treatment is meant to be free at government facilities due in part by support from international donors.

Approximately half of the participants in our survey indicated that it takes them over an hour to reach a health facility. Forty-six and 39% of the respondents waited one and two days respectively before seeking medical care for illness. A similar delay to seek care was also reported from Bangladesh (Ahmed at al., 2009). A previous KAP survey in a different area of Guinea found that seeking treatment in a formal health facility was dependent on socioeconomic status (Ruberto et al., 2014). Initial treatment at home and waiting for more than 24 hours was also seen in Equatorial Guinea (Romay-Barja et al., 2015). Logistical obstacles and reliance on traditional remedies have been cited as major barriers to malaria treatment in a systematic review (Maslove, Mnyusiwalla et al. 2009).

Almost all participants in our survey indicated that malaria is a serious illness in adults and in children. This was similar to what was reported from Swaziland (Hlongwana et al., 2009). We saw a large percentage of our respondents preferring injectable malaria treatment as they considered it to be stronger and faster in action. This is another cause for concern since injections can cause complications such as infections and abscesses as well as necessitate proper disposal of sharps. This curious practice of injectable malaria treatment has previously been reported from Guinea by a much earlier study, showing the persistence of such treatment behavior across more than two decades (Dabis, Breman et al. 1989).

### Information source

Radio was the most cited and trusted source of information in our survey. Television was cited as the most common source of malaria information in the much richer Cabo Verde (DePina et al., 2019). Media was also cited as the most common source of information on malaria behavioral change communication in Nigeria (Kilian et al., 2016).

## Conclusion

Guinea was struggling with reducing malaria mortality even before the arrival of COVID-19. The current target of achieving pre-elimination by 2022 will become even more challenging in this time when national malaria control programs worldwide are predicting serious setbacks from interruption of efforts. Our study documents a detailed collage of knowledge, attitudes and practices among members as they relate to malaria in a rural Guinean community with a high malaria burden. We saw a community where most caregivers of children under five were women with very little formal education. Despite this lack of education, they knew the causes of malaria and diarrhea, and they were taking their children for preventive care to local health posts.

Malaria and diarrhea were considered serious illnesses and were treated at both formal, informal, and traditional healthcare providers. We saw high usage of bednets and their contribution to reducing malaria. We have identified gaps in current knowledge as well as suggested how to target those gaps. We have shown what practices have been effective in reducing the burden of malaria. Comparing our findings with those of others from Guinea as well as from other countries makes it clear that effective interventions to control and prevent malaria will need a detailed understanding of local facts. Our study adds valuable information for policy makers and practitioners who design and implement malaria control and prevention measures in Guinea.

## Data Availability

The datasets used in this study are available from the corresponding author on reasonable request.

## Declarations

### Ethics approval and consent to participate

Informed verbal consent was obtained from each survey participant prior to administering the survey. Ethics approval for the study was obtained from the Guinea National Ethics Committee for Health Research (96/CNERS/16).

### Consent for publication

### Not applicable

### Availability of data and materials

All data generated or analyzed during this study are included in this published article [and its supplementary information files].

## Authors’ contributions

EH analyzed the data and drafted the manuscript. BK designed the study and acquired the data. AS conceptualized and supervised the study. AS analyzed the data and substantially revised the manuscript. NR designed the study, interpreted the data and substantially revised the manuscript. All authors read and approved the final manuscript.

## Acknowledgements

The authors wish to thank Dr. Kathryn H. Jacobsen of George Mason University for her invaluable assistance developing our survey instrument.

